# Cardiorespiratory Dynamics During Prone Positioning for Severe Ards: A Retrospective Cohort Study

**DOI:** 10.1101/2024.01.30.24301935

**Authors:** Andrew Barros, Seung Wook Lee, J. Randall Moorman

**Affiliations:** Center for Advanced Medical Analytics (CAMA), School of Medicine, University of Virginia, Charlottesville, VA; Division of Pulmonary and Critical Care Medicine, Department of Medicine, School of Medicine, University of Virginia, Charlottesville, VA; Division of Cardiovascular Medicine, Department of Medicine, School of Medicine, University of Virginia, Charlottesville, VA

**Author notes:** These authors contributed equally to this work (Andrew Barros, Seung Wook Lee).

## Abstract

**Objectives:** To elucidate the changes in cardiorespiratory dynamics during neuromuscular blockade and prone positioning and determine the associations between changes in cardiorespiratory dynamics following prone positioning and mortality.

**Design:** Single center retrospective cohort study of patients admitted to the medical ICU between June 1, 2020 and September 1, 2022 who received prone positioning while mechanically ventilated.

**Results:** Our final cohort consisted of 136 patients. Prone position was associated with an improvement in A-a gradient of 113 mmHg (95% CrI 78 – 149) between the pre-proning values and 10 hours post proning. Norepinephrine dose did not significantly change before and after prone positioning (Estimated difference: 0.04 mcg/min 95% CrI -1.00 – 1.07). For the outcome of 7-d mortality, there was a high probability that the baseline factors of increasing age, male sex, and higher baseline A-a gradient were associated with increased risk of death. Increased total vasopressor requirement and increased in PCO2 were associated with worse prognosis while a decrease in instantaneous heart rate and a decrease in heart rate variability were associated with improved prognosis.

**Conclusion:** The immediate changes in prone positioning primarily impact respiratory physiology, with limited influence on circulatory parameters. Predictors of short-term mortality after prone positioning include both respiratory and cardiovascular parameters suggesting that extrapulmonary effects, such as improvement in right ventricular heart function, might also contribute to the benefit of prone positioning.

## INTRODUCTION

Acute respiratory distress syndrome (ARDS) is a grave respiratory ailment characterized by a high mortality rate and significant comorbidity in survivors. It primarily involves increased permeability of the alveolar-capillary barrier, resulting in diminished oxygen exchange, reduced lung compliance, and pulmonary edema. The hospital mortality rate for ARDS varies between 35% and 45%, depending on disease severity, with more severe cases having higher mortality rates^1^. Consequently, the timely recognition and appropriate treatment of ARDS are paramount in reducing mortality and enhancing clinical outcomes.

While patients with mild ARDS and adequate oxygenation can be managed conservatively, those with moderate to severe ARDS typically necessitate mechanical ventilation. In cases of severe ARDS where conventional treatments prove ineffective, clinicians may consider prone positioning as an alternative strategy. Prone positioning, initially shown to improve oxygenation in ARDS patients in the 1970s^2,3^ and reduce mortality in a randomized trial^4^, is now frequently employed as a rescue therapy for severe ARDS patients unresponsive to traditional treatments. The use of this maneuver witnessed a surge during the COVID-19 pandemic^5^, as a substantial portion of COVID-19 patients hospitalized for acute respiratory failure progressed to moderate to severe ARDS, requiring mechanical ventilation. The rapid progression of the disease, coupled with its high prevalence, placed unprecedented strain on healthcare systems, leading to shortages of intensive care unit (ICU) capacity and mechanical ventilators. This scarcity of resources escalated the demand for alternative respiratory management strategies, such as prone positioning, to alleviate the burden.

In addition to its well-documented benefits in oxygenation^6,7^, there is growing evidence supporting the mortality-reducing effects of prone positioning. A meta-analysis involving 1,867 patients reported a 16% reduction in mortality in patients with severe hypoxemia (PaO2/FiO2 < 100 mmHg) (risk ratio [RR] 0.84, 95% confidence interval 0.74 - 0.96, p = 0.01)^8^, and similar results have been reported in other meta-analyses^9–11^. Furthermore, the PROSEVA trial demonstrated a reduction in 90-day mortality (hazard ratio 0.44, 95% CI 0.29 to 0.67) with early, prolonged prone positioning in mechanically ventilated patients with severe ARDS^4^.

While studies have extensively documented the physiological effects of prone positioning on oxygenation, such as improved lung perfusion^12^ and reduced differences in ventral-dorsal transpulmonary pressure^13^, it is essential to recognize that the effects of this maneuver extend beyond the respiratory system. Prone positioning has been found to exert extrapulmonary hemodynamic effects, including an increase in cardiac output^14^ and improvements in right ventricular volume-pressure overload^15^. Paired with the observations that right ventricular dysfunction is both common in ARDS^16^ and associated with increased mortality^17^ the observed benefits of prone positioning may derive from a composite of both respiratory and non-respiratory effects.

In this study, we aim to investigate the cardiopulmonary characteristics of a cohort of patients who underwent prone positioning for severe ARDS. Specifically, our objectives are to 1) elucidate the changes in cardiorespiratory dynamics during neuromuscular blockade and prone positioning and 2) determine the associations between changes in cardiorespiratory dynamics following prone positioning and mortality.

## METHODS

### Overall Study Design and Data Collection

This retrospective, observational cohort study of adults with severe ARDS was conducted at a single academic medical center. We used our enterprise data warehouse to identify patients admitted to the medical ICU service between June 1, 2020 and September 1, 2022 who had any nursing documentation of prone positioning. These encounters were screened by an investigator (SWL) who confirmed the time and indication for prone positioning from documentation. The IRB of The University of Virginia gave ethical approval for this work (HSR-IRB 22152). This work follows the Strengthening the Reporting of Observational Studies in Epidemiology (STROBE) guidelines^18^.

### Analysis: Changes at the time of NMB and Prone Positioning

#### Data Collection

For the outcome of neuromuscular blockade, we included data from ten hours before NMB administration until 30 minutes before NMB administration and 30 minutes after NMB administration until one hour before prone positioning. For the outcome of prone positioning, we included data from after NMB administration until 1 hour before prone positioning and 1 hour after prone positioning until 8 hours after prone positioning.

#### Predictors

We extracted demographics (age at admission, sex), vital signs, laboratory measures (lactate, blood gases), administered medications (vasopressors and neuromuscular blockers), and cardiorespiratory features.

#### Outcome

Our primary outcome was an indicator variable for if a time stamp occurred before or after our target event (NMB or prone positioning)

#### Preprocessing

We used a last observation carried forward approach to create observations every 15 minutes during the observation periods. We calculated the total norepinephrine equivalent dose and estimated a-A gradient.

#### Statistical Methods

We used a Bayesian mixed effects generalized additive model with a logit link (i.e. logistic regression), P-splines restricted to a basis dimension of 4 for the fixed effects, a random effect for the subject, and weakly informative priors (i.e. Normal (0,2.5) on the logit scale). We estimated the model using Hamiltonian Monte Carlo methods with 8 chains, 4000 total iterations, and 2000 warmup iterations.

### Analysis: Predictors of Mortality after Prone Positioning

#### Data Collection

We considered six windows around the proning event: Baseline (four hours before to 1 hour before proning), one-hour (one hour after to two hours after proning), two-hour (two hours after to three hours after), four-hour (three hours after to five hours after), six-hour (five hours after to seven hours after), and eight-hour (7 hours after to 9 hours after). Observations in each window were averaged to produce a single summary value.

#### Predictors

Our predictors were the same as the prior analysis.

#### Outcome

Our primary outcome was 7-day mortality or discharge to hospice.

#### Preprocessing

For each window we included the baseline features and the difference between the baseline and that time window.

#### Statistical Methods

We used a Bayesian generalized linear model with a logit link and weakly informative priors (Bayesian Logistic Regression). We estimated the model using Hamiltonian Monte Carlo methods with 8 chains, 2000 total iterations, and 1000 warmup iterations.

### Statistical Methods

For both analyses, we used a preprocessing pipeline with Python and Pandas. The analysis was conducted in R (4.3.0) using BRMS^19^ and Stan^20^. We used the maximum a posteriori (MAP) estimates for point estimates and 95% equal tailed intervals for credible intervals. The largest percentage of the probability distribution on either side of the null result was defined as the probability of direction. A fixed seed was used for all analyses for reproducibility.

## RESULTS

We initially included 190 patients but subsequently excluded 54: 19 did not have recorded cardiorespiratory dynamics measures, 5 received neuromuscular blockade prior to admission, and 30 had less than one hour between neuromuscular blockade and prone positioning (eFigure 1). Our final analysis cohort consisted of 136 patients (Table 1).

**Table 1:** Cohort Demographics.

| Characteristic | N = 136<sup>1</sup> |
| --- | --- |
| Age, years | 62 (53, 69) |
| Male Sex | 51 (38%) |
| In Hospital Mortality | 84 (62%) |
| 7 Day Mortality | 31 (23%) |
| 30 Day Mortality | 73 (54%) |
| Minimum Pre-Prone PF Ratio | 74 (65, 88) |
| Minimum Pre-Prone SF Ratio | 91 (86, 97) |
| Maximum Pre-Prone A-a Gradient | 560 (532, 577) |

<sup>1</sup> Median (IQR); n (%)

Figure 1 shows the unadjusted distribution of features for both neuromuscular blockade and prone positioning. In this figure, a plausible physiologic effect would be displayed as a smooth gradient of color from blue to red and greater odds (red) is associated with a greater chance than a value in that range was observed after the event of interest. The largest effect is in the oxygenation parameters (A-a gradient, S:F ratio, and P:F ratio) after prone positioning (Figure 1c) with little effect seen on circulatory parameters (Total norepinephrine dose, mean arterial pressure). This is consistent with a larger effect on respiration than circulation. For the whole cohort, prone position was associated with an improvement in A-a gradient of 113 mmHg (95% CrI 78 – 149) between the pre-proning values and 10 hours post proning (Figure 2). Norepinephrine dose, on the other hand, did not significantly change before and after prone positioning (Estimated difference: 0.04 mcg/min 95% CrI -1.00 – 1.07).

**Figure 1.**
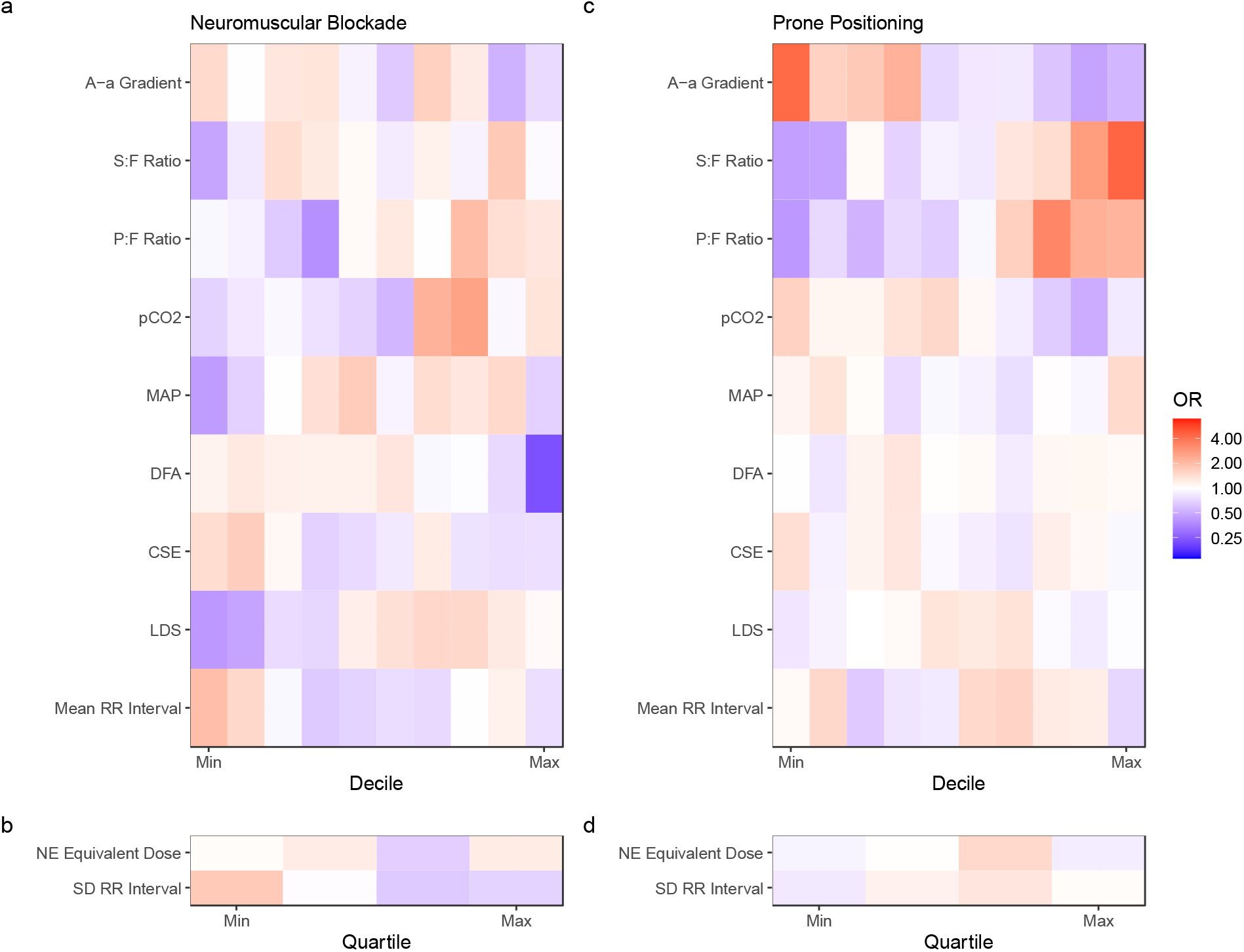
Changes in cardiorespiratory parameters after neuromuscular blockade and prone positioning. Odds ratio (color) of the post neuromuscular blockade (panels a and b) or prone positioning (panels c and d) state as a function of the quartile (panels b and d) or decile (panels a and c) for the parameters listed along the left. For example, the red color in the upper left box of panel C signifies that the lowest decile of A-a gradient was four times more likely to be after prone positioning than before prone positioning. Note the top row in panel (c) shows monotonic relationship of A-a gradient with lower values having a high likelihood of being observed after proning (red colors) and high values have a low likelihood of being observed after proning (blue colors). In contrast, A-a gradient in panel (a) has an inconsistent trend.

**Figure 2.**
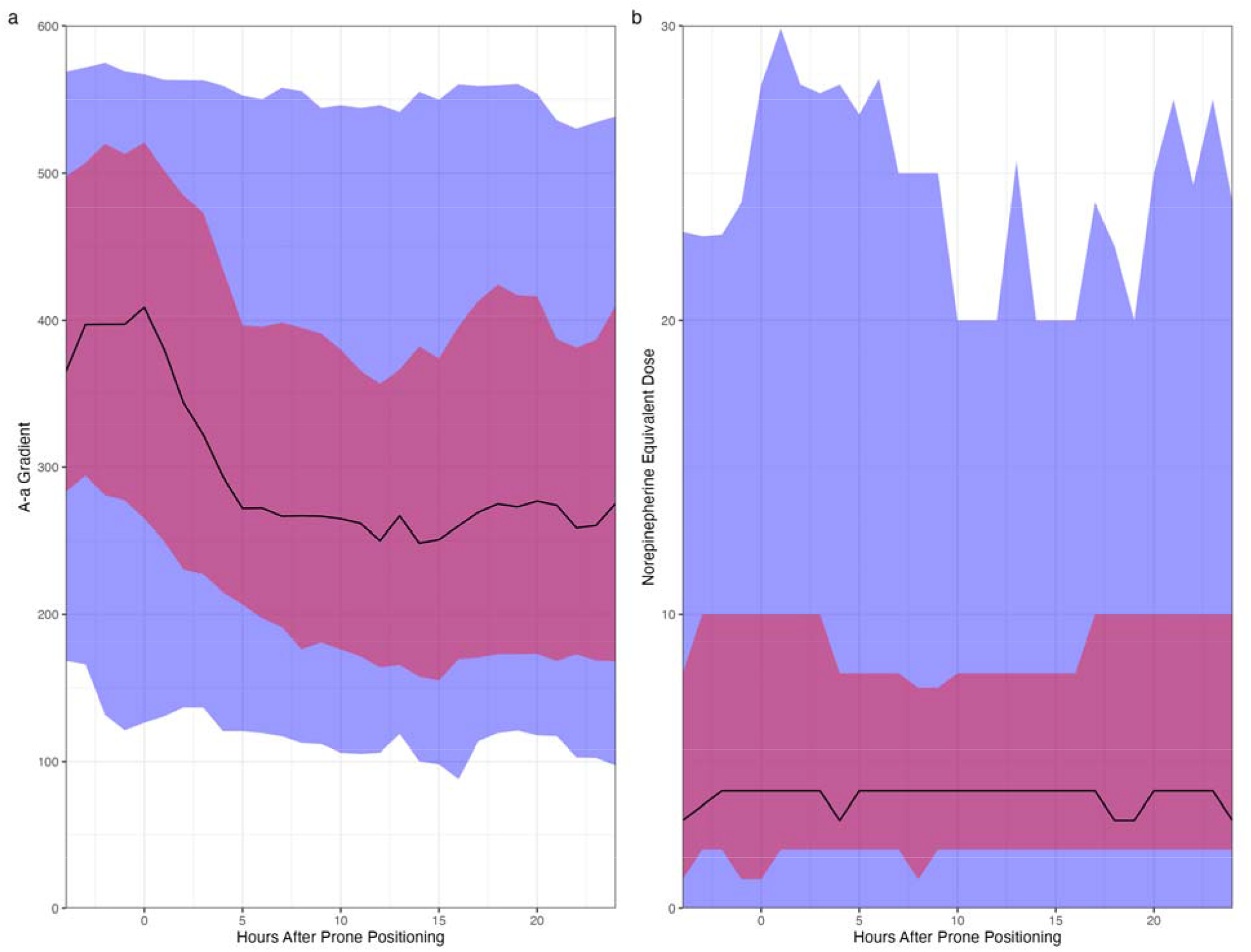
The distributions of observed values for (a) A-a gradient and (b) norepinephrine equivalent dose for four hours before and 24 hours after prone positioning. The black line represents the median value while the blue and red shaded regions represent the 5^th^ to 95^th^ and 25^th^ to 75^th^ percentile values respectively. Observations were aligned to 15-minute boundaries and carried forward until a change was observed.

When we examine the combined effects (Figure 3), neuromuscular blockade was estimated to have a moderate effect on the coefficient of sample entropy, average instantaneous heart rate, and PCO2 while A-a gradient, detrended fluctuation analysis of the heart rate, PCO2, total norepinephrine dose, mean arterial blood pressure, coefficient of sample entropy, local dynamics score, and heart rate were estimated to have small effects. This is consistent with broad cardiovascular and respiratory effects of neuromuscular blockade. For prone positioning, only A-a gradient and PCO2 were estimated to change after prone positioning. This is consistent with the primary effect of prone positioning primarily on the respiratory system.

**Figure 3.**
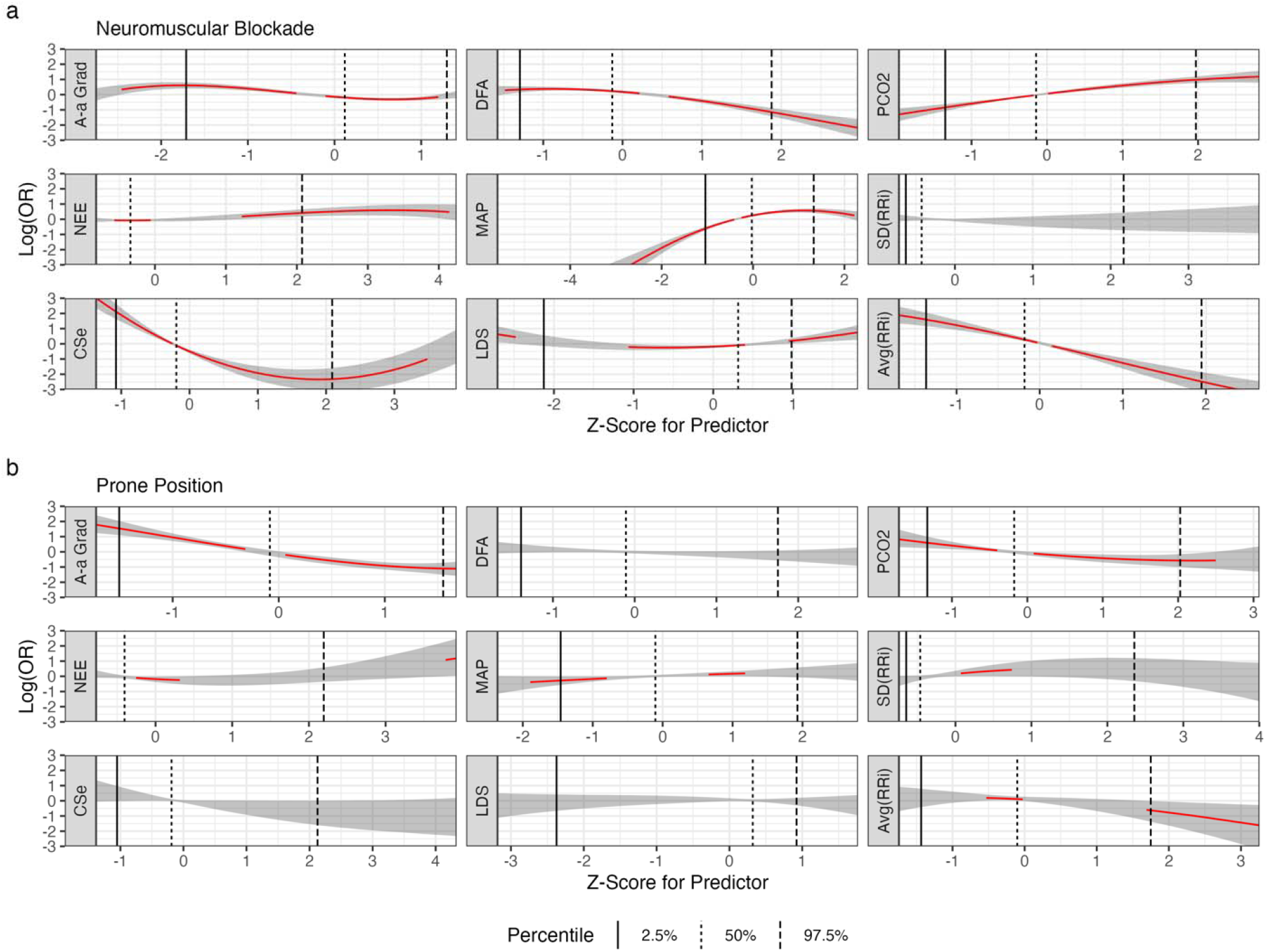
Estimated effects for cardiorespiratory predictors before and after (a) neuromuscular blockade and (b) prone positioning. The red line represents the MAP estimate from the posterior predictive and is omitted when the 95% highest density posterior interval contains the null effect. The grey band represents the 95% highest density posterior interval. The vertical lines represent the percentiles of observed values. Panel A shows that the effects of neuromuscular blockade are diverse with observed impact on ventilation (A-a gradient and pCO2), hemodynamics (norepinephrine equivalent dose and mean arterial pressure), and heart rate dynamics (coefficient of sample entropy and instantaneous heart rate). In contrast, Panel B shows that the dominant effect of prone positioning is on ventilation with modest effects on hemodynamics and nearly no effect on heart rate dynamics.

For the outcome of 7-d mortality, there was a high probability that the baseline factors of increasing age, male sex, and higher baseline A-a gradient were associated with increased risk of death (Table 2). Worsening shock as demonstrated by increased total vasopressor requirement was associated with worse prognosis after the first hour. In contrast, an increase in PCO2 was associated with worse prognosis at the first hour with decreasing effect seen over time. A decrease in instantaneous heart rate, corresponding to an increase in mean RR interval, and a decrease in heart rate variability, corresponding to a decrease in standard deviation of the RR interval, were associated with improved prognosis. A comparison of selected predictors and their association with the events of neuromuscular blockage and prone positioning are shown in Figure 4.

**Table 2:** Model Estimates.

| Variable | 1h | 2h | 4h | 6h | 8h |
| --- | --- | --- | --- | --- | --- |
| Age/Decade | **2.48 [1.25-5.26, 99.7%]** | **2.30 [1.21-4.79, 99.6%]** | **2.20 [1.21-4.25, 99.5%]** | **1.83 [1.06-3.43, 98.3%]** | **1.98 [1.10-3.84, 99.0%]** |
| Male Sex | 2.19 [0.55-9.02, 86.1%] | 1.81 [0.45-7.60, 80.2%] | 2.68 [0.68-11.13, 92.1%] | 2.90 [0.73-11.97, 93.8%] | 2.04 [0.49-8.64, 84.2%] |
| AA Gradient | **3.58 [1.54-9.56, 99.9%]** | **3.80 [1.62-10.20, 99.9%]** | **4.03 [1.72-10.69, 100.0%]** | **4.37 [1.72-11.91, 100.0%]** | **3.27 [1.26-8.81, 99.4%]** |
| AA Gradient Change | 0.99 [0.30-3.30, 51.0%] | 0.89 [0.29-2.68, 57.9%] | 1.94 [0.69-5.89, 88.7%] | 2.35 [0.80-7.01, 94.4%] | 1.76 [0.61-5.20, 85.3%] |
| CSe | 0.40 [0.04-2.83, 80.3%] | 0.43 [0.04-3.19, 76.9%] | 0.32 [0.03-2.74, 83.5%] | 0.25 [0.02-2.33, 87.1%] | 0.31 [0.03-2.46, 84.5%] |
| CSe Change | 0.53 [0.03-11.30, 66.0%] | 1.34 [0.07-29.78, 57.7%] | 0.42 [0.03-5.85, 73.8%] | 0.55 [0.04-7.96, 67.7%] | 0.50 [0.09-2.70, 78.0%] |
| NE Total Dose | 0.61 [0.20-1.65, 81.3%] | 0.87 [0.33-2.15, 60.2%] | 1.04 [0.39-2.58, 55.0%] | 0.92 [0.33-2.25, 53.5%] | 0.84 [0.30-2.15, 63.3%] |
| NE Total Dose Change | 1.06 [0.17-6.68, 52.0%] | **2.78 [0.83-9.91, 95.2%]** | **3.72 [1.12-13.72, 98.5%]** | 2.51 [0.72-8.98, 92.2%] | 1.71 [0.40-7.42, 76.6%] |
| PCO2 | 1.16 [0.55-2.45, 65.5%] | 1.30 [0.62-2.75, 75.2%] | 1.33 [0.62-2.82, 77.9%] | 0.98 [0.44-2.16, 50.8%] | 1.03 [0.42-2.44, 53.8%] |
| PCO2 Change | **7.37 [1.92-38.06, 99.9%]** | **6.54 [2.01-27.51, 100.0%]** | **2.54 [0.99-7.07, 97.3%]** | 1.11 [0.54-2.36, 59.9%] | 1.16 [0.49-2.81, 63.1%] |
| Avg(RRi) | 0.53 [0.09-2.87, 75.7%] | 0.55 [0.09-3.10, 75.5%] | 0.63 [0.10-3.43, 69.6%] | 0.40 [0.06-2.44, 82.8%] | 0.43 [0.07-2.45, 82.1%] |
| Avg(RRi) Diff | **0.04 [0.00-0.47, 99.5%]** | **0.13 [0.01-1.15, 96.8%]** | 0.39 [0.05-2.95, 81.2%] | 0.43 [0.06-3.40, 79.2%] | 0.37 [0.06-1.98, 87.3%] |
| SD(RRi) | 1.79 [0.33-9.43, 74.8%] | 1.79 [0.33-9.23, 76.1%] | 1.58 [0.28-8.54, 70.7%] | 2.74 [0.42-17.09, 86.0%] | 2.42 [0.36-15.49, 82.2%] |
| SD(RRi) Change | **14.24 [1.92-123.29, 99.5%]** | **5.23 [0.93-41.28, 97.0%]** | 3.70 [0.61-25.15, 91.9%] | **4.50 [0.81-28.63, 95.3%]** | 3.15 [0.65-18.39, 91.9%] |

**Figure 4.**
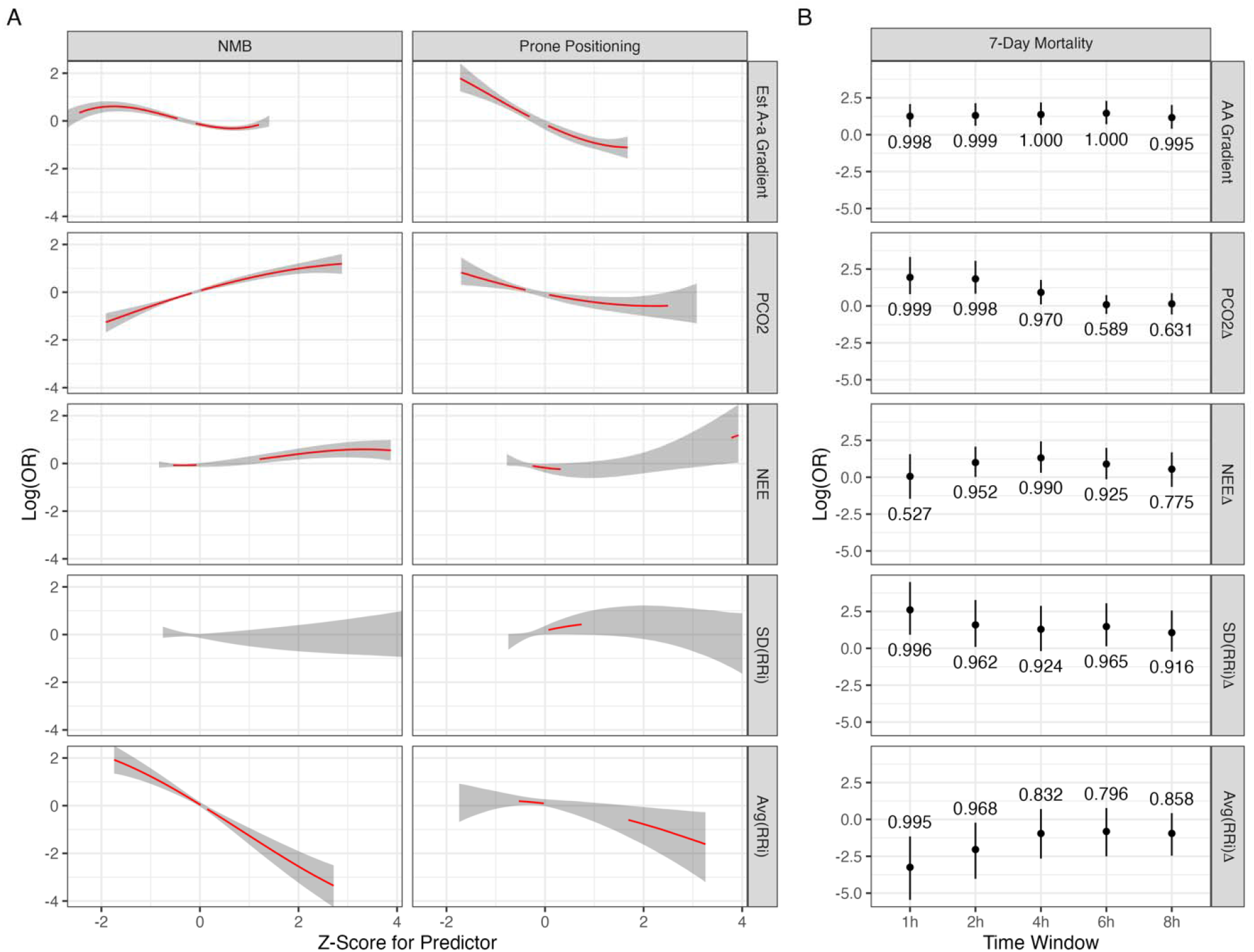
This graph shows that the effect of prone positioning and neuromuscular blockade (Panel A) for a subset of predictors and the association of those predictors with 7-day mortality (Panel B). In panel A, the mean value from the posterior predictive is shown in red and the line is omitted when the 95% credible interval includes no effect. The grey band represents the 95% credible interval for the posterior predictive. Note that many of the estimated effects are small with little variation over the range of commonly observed values (Z -2 to 2). However, the estimated A-a gradient and the instantaneous heart rate (Average RR interval) have large estimated effects for the events of prone positioning and neuromuscular blockade respectively. In panel B, the adjusted association of selected predictors after prone positioning with 7-day mortality. The closed circle is the mean posterior parameter estimate and bars represent the 95% credible interval. The probability that the parameter is greater or less than zero is above the line. The first row shows the estimated effect of the baseline A-a gradient in a joint model with the changes at each time point. The estimated effect is consistent with a high probability of direction. In contrast, the second line shows the effect of the change in pCO2 with immediate increases associated with increased mortality (and immediate decreases associated with decreased mortality). The effect is smaller and less certain as time progresses after prone positioning.

## DISCUSSION

In summary, the phenotype of improved prognosis after proning was associated with decreased vasopressor requirement, improved ventilation, decreased heart rate, and decreased heart rate variability. One explanation of these findings is improved right ventricular function resulting from improving oxygenation and ventilation due to prone positioning.

Our analysis supports previous evidence regarding the beneficial effects of prone positioning on oxygenation and ventilation. These improvements are consistent with the primary physiological effect of prone positioning on the respiratory system, which includes enhanced lung perfusion and decreased ventilation-perfusion mismatch. Additionally, our analysis did not show large immediate changes in cardiovascular parameters after prone positioning. The findings of this study shed light on the complex interplay of physiologic factors resulting from the use of prone positioning as a therapeutic strategy for severe ARDS.

Interestingly, neuromuscular blockade was associated with changes in several cardiorespiratory predictors, including A-a gradient, heart rate variability, and PCO2. This observation underscores the broader physiological effects of neuromuscular blockade beyond muscle relaxation. However, it is worth noting that only a few of these changes had a significant clinical impact, emphasizing the need for careful consideration of the risks and benefits of neuromuscular blockade in ARDS management.

Furthermore, our analysis of predictors of short-term mortality following prone positioning revealed several key insights. Increasing age and higher baseline A-a gradient were associated with an elevated risk of mortality consistent with previously published work. Worsening shock, as evidenced by increased total vasopressor requirements, was predictive of worse prognosis after the first hour post-proning. Conversely, an increase in PCO2 was initially associated with a worse prognosis, but its effect diminished over time. This suggests that the dynamics of carbon dioxide levels may have complex implications for ARDS outcomes.

The observation that a decrease in instantaneous heart rate and heart rate variability was associated with improved prognosis raises intriguing questions about the role of cardiac function in ARDS management. It is plausible that improved right ventricular function, resulting from enhanced oxygenation and ventilation due to prone positioning, contributes to the observed changes in heart rate parameters.

## CONCLUSION

In conclusion, this study contributes valuable insights into the effects of prone positioning and neuromuscular blockade in severe ARDS management. The immediate changes in prone positioning primarily impact respiratory physiology, with limited influence on circulatory parameters. In contrast, neuromuscular blockade exhibited a more nuanced effect with broad effects on respiratory and cardiovascular physiology. Predictors of short-term mortality after prone positioning include both respiratory and cardiovascular parameters suggesting that extrapulmonary effects, such as improvement in right ventricular heart function, might also contribute to the benefit of prone positioning.

## Data Availability

All data produced in the present study are available upon reasonable request to the authors

## DATA AVAILABILITY

All data produced in the present study are available upon reasonable request to the authors.

## ABBREVIATIONS

ARDS: Acute respiratory distress syndrome
ICU: Intensive care unit
FiO2: Fraction of inspired oxygen
PaO2: Partial pressure of oxygen in the arterial blood
SpO2: Oxygen saturation measured in pulse oximetry
NMB: Neuromuscular blockade
A-a gradient: Alveolar to Arterial Oxygen Gradient
PF Ratio: Ratio of partial pressure of oxygen in arterial blood (PaO2) to the Fraction of inspiratory oxygen concentration (FiO2)
SF Ratio: Ratio of Oxygen saturation (Spo2) to the Fraction of inspiratory oxygen concentration (FiO2)
pCO2: Partial Pressure of carbon dioxide
MAP: Mean Arterial Pressure
DFA: Detrended Fluctuation Analysis of the Heart Rate
CSE: Coefficient of Sample Entropy
LDS: Local Dynamics Score

## CONTRIBUTIONS

Conceptualization: AJB, JRM

Methodology: AJB, SWL, JRM

Software: AJB, SWL

Validation: AJB, JRM

Formal analysis: AJB, SWL

Investigation: AJB, SWL

Resources: JRM

Data Curation: AJB, SWL

Writing - Original Draft: AJB, SWL

Writing - Review & Editing: AJB, SWL, JRM

Visualization: AJB

Supervision: AJB, JRM

Project administration: AJB

Funding acquisition: JRM

## FIGURES

**eFigure 1.**
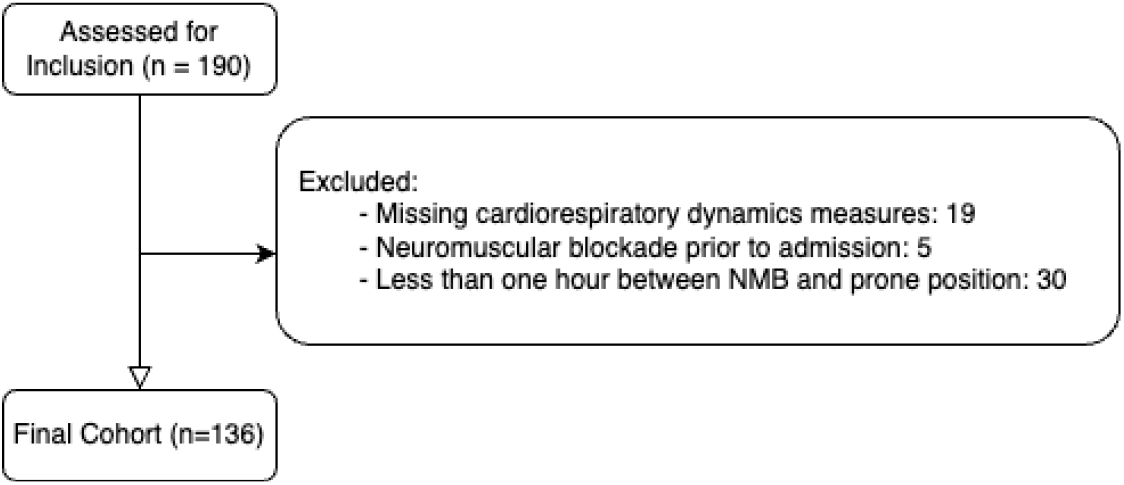

